# KRAS Inhibition in Pancreatic Cancer: Pooled Efficacy and Safety Signals from Early-Phase Studies

**DOI:** 10.64898/2025.12.03.25341547

**Authors:** Kathlen Oliveira Martins Tiede, Maria Fernanda Teixeira, Mariana Moura, Ohana Chiaini, Fernando Moura, Mohamad Bassam Sonbol, Mitesh Borad, Tanios Bekaii-Saab, Pedro Luiz Serrano Uson Junior

## Abstract

Pancreatic ductal adenocarcinoma (PDAC) remains one of the most lethal cancers, driven by KRAS mutations long deemed “undruggable”. We conducted a meta-analysis of five early phase cohorts (n=301) that evaluated KRAS-targeted therapies in patients with PDAC. The pooled objective response rate was 28% (95% CI 22–35%), indicating promising activity in refractory PDAC, with consistent estimates across the studies (I^2^=0%). Gastrointestinal toxicities were common (diarrhea, 47%; nausea, 43%). These findings validate direct KRAS inhibition as a breakthrough concept in PDAC but are tempered by modest durability, risk of bias, and limitations of early phase designs, underscoring the need for biomarker-guided, rigorously designed clinical trials.

## Main text

Pancreatic ductal adenocarcinoma (PDAC) is characterized by its aggressiveness and dismal prognosis, particularly in the metastatic setting(1). Standard systemic therapy consists of multi-agent chemotherapy, such as fluorouracil-based combinations or gemcitabine plus nab-paclitaxel (2),(3). While these regimens modestly prolong survival, toxicity is often related to dose reductions or early discontinuation (4),(5). Activating mutations in RAS, most notably KRAS, are present in approximately 90% of cases and are the major oncogenic drivers in PDAC (6), (7). Despite decades of recognition as a critical oncogene, KRAS was long considered “undruggable” because of its high affinity for GTP/GDP and lack of accessible binding pockets, limiting progress to indirect pathway blockade with limited efficacy (8). However, recent advances have enabled the development of small molecules that directly disrupt KRAS signaling, representing a long-sought breakthrough (9), (10).

Several agents have been engineered, including adagrasib, sotorasib, and daraxonrasib (a covalent inhibitor that irreversibly locks KRAS in its inactive conformation)(11, 12), (13, 14) (15), (16). The siG12D-LODER trial, which used a biodegradable polymeric matrix to deliver KRAS-directed therapy locally, was not included in our analysis as it combined the intervention with chemotherapy in the locally advanced setting, differing from the monotherapy trials of small-molecule inhibitors (17). In patients with PDAC, these approaches remain largely in the early stages of development, with only limited clinical reports and conference presentations. To synthesize the available evidence, we performed a systematic review and meta-analysis of KRAS-targeted therapy. The primary endpoint was the objective response rate (ORR), with safety assessed by the frequency of common adverse events. Five non-randomized phase I/II cohorts encompassing 301 patients with metastatic or locally advanced PDAC treated with KRAS inhibitors in the second-line or beyond met the inclusion criteria [**Table 1**] (PRISMA flow diagram and study characteristics are shown in the Supplementary Appendix). Of the 301 patients included in the study, only 205 were evaluated for the objective response rate (ORR). The remaining patients were excluded from the analysis due to the absence of adequate imaging assessments, loss to follow-up, or other reasons that prevented response evaluation.

**Table 1:** Characteristics of the included studies evaluating KRAS inhibitors in pancreatic ductal adenocarcinoma (PDAC). OR = Objective Response, DCR = Disease Control Rate, DoR = Duration of Response, PFS = Progression-Free Survival, TRAEs = treatment-related adverse events, TTM = time to metastasis, DLT= dose-limiting toxicity, QD = once daily, PR= partial response, NR= not reported.

| Study | Country | Study Phase | KRAS inhibitor | Number of patients | Age | Mutation type | Metastatic or locally advanced | Dose | Follow-up | Outcomes reported | ORR | DCR | DoR | PFS |
| --- | --- | --- | --- | --- | --- | --- | --- | --- | --- | --- | --- | --- | --- | --- |
| Stricker, 2023 | Multinational | Phase II | Sotorasib | 38 | 65.5 (45-81) | KRAS p. G12C | Metastatic | 960mg | 18 months | ORR, DCR, DOR, PFS, OS, TRAEs | 21% | 84% | 5.7 mo (1.6 - NA) | 4 mo (2.8 -5.6) |
| Xinghao Ai, 2025 | China | Phase I/IIa | GFH375 | 32 | 59.5 | KRAS p. G12D | Locally Advanced and Metastatic | 100-900mg QD e 300mg BD | NR | ORR, TRAEs | 43% | 100% | NR | NR |
| Garrido-Laguna, 2025 | United States | Phase I | Daraxonrasib | 126 | 64 (30-86) | KRAS p.G12X and RAS mutations | Metastatic | 160-300mg QD | ≥14 weeks | ORR, DCR, PFS, OS, TRAEs, ctDNA clearance | 27% | 90% | NR | G12X: 8.5mo (5.3–11.7)<br>RAS mutations: 7.6mo (5.9–11.1) |
| Spira, 2025 | United States | Phase I | Zoldonrasib | 104 | 65 (30-86) | KRAS p. G12D | Metastatic and Locally Advanced | 150 - 1200mg QD | ≥14 weeks | ORR, DCR, ctDNA clearance, TRAE | 30% | 80% | NR | NR |
| Bekaii-Saab, 2023 | United States and Puerto Rico | Phase I/II | Adagrasib | 21 | 65 (21-89) | KRAS p. G12C | Metastatic and Locally Advanced | 600mg BID | 16.8 months | ORR, DCR, PFS, OS, TRAEs | 33% | 81% | NR | 5.4 mo (3.9-8.2) |

The pooled ORR across studies was 28%, reflecting measurable antitumor activity in otherwise treatment-refractory disease (18). Although KRAS-targeted agents have demonstrated activity across classes, the magnitude of their benefit remains modest compared to that in other malignancies. In pancreatic cancer, particularly KRAS G12D-mutated tumors, even incremental improvements are of clinical relevance, given the historically poor prognosis. Current studies remain in the early stages of development, with immature survival outcomes; however, these data collectively provide proof-of-concept that direct KRAS inhibition is feasible in PDAC.

The efficacy estimates were consistent, with no statistical heterogeneity (I^2^ = 0%). The narrow prediction interval, which was closely aligned with the confidence interval, reinforced the robustness of the pooled ORR and suggested that future trials are likely to yield comparable results [**Figure 1**]. Meta-regression showed no association between study size and overall response rate. Although smaller trials had wider confidence intervals, the efficacy estimates were consistent across the studies [**Figure 2**].

**Figure 1.**
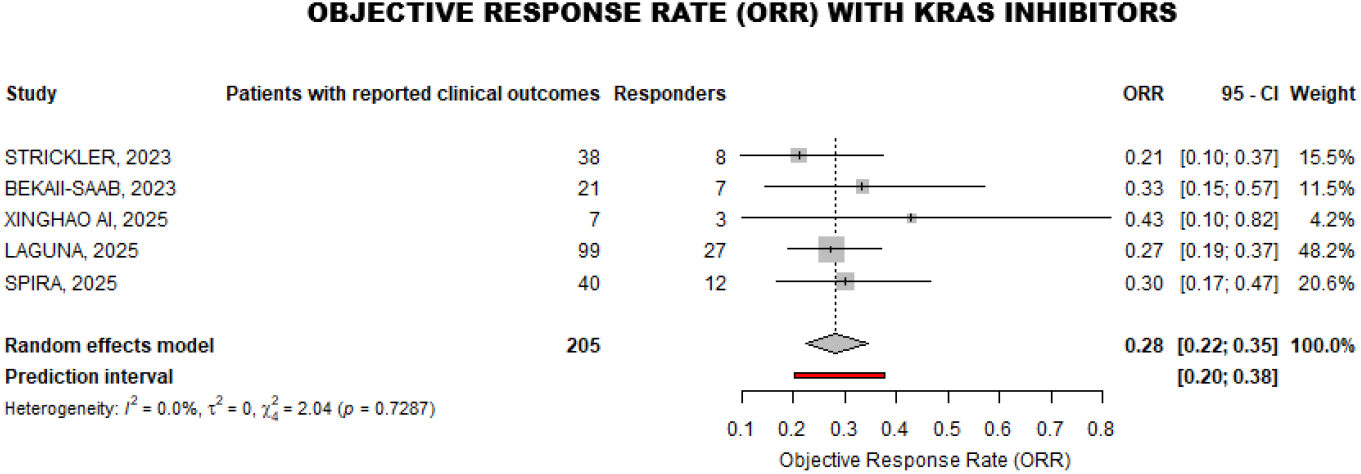
Objective response rate (ORR) with KRAS inhibitors. The pooled ORR across five trials (205 evaluable patients out of 301 enrolled) was 28% (95% CI, 22–35%), with narrow prediction intervals and no significant heterogeneity (I^2^ = 0%).

**Figure 2:**
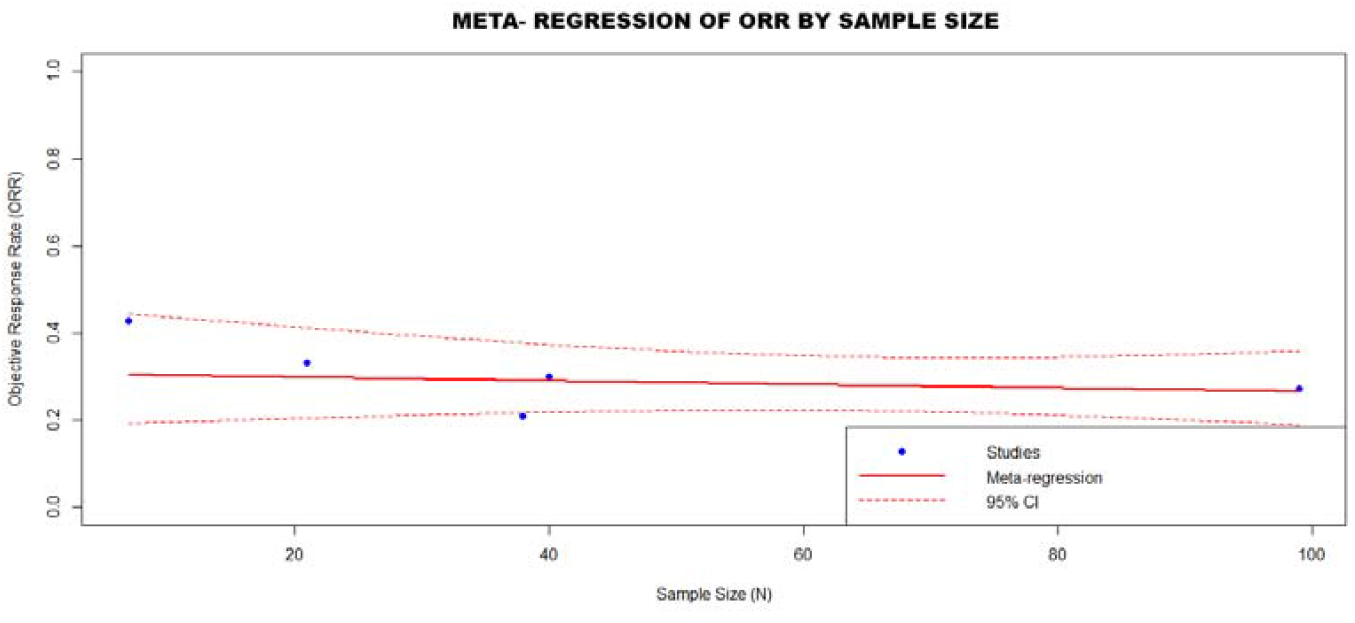
Meta-regression of ORR by sample size. No association was observed between the study sample size and objective response rate (ORR).

Gastrointestinal toxicities were common, with diarrhea (47%) and nausea (43%) affecting more than one-third of the patients [**Figure 3**]. Confidence and prediction intervals were wide, and leave-one-out analyses identified that no single study explained the heterogeneity effects, which reinforces the variability among the included studies [Supplementary Appendix]. Larger trials with standardized safety reporting are required to better define the safety profile of KRAS inhibitors.

**Figure 3:**
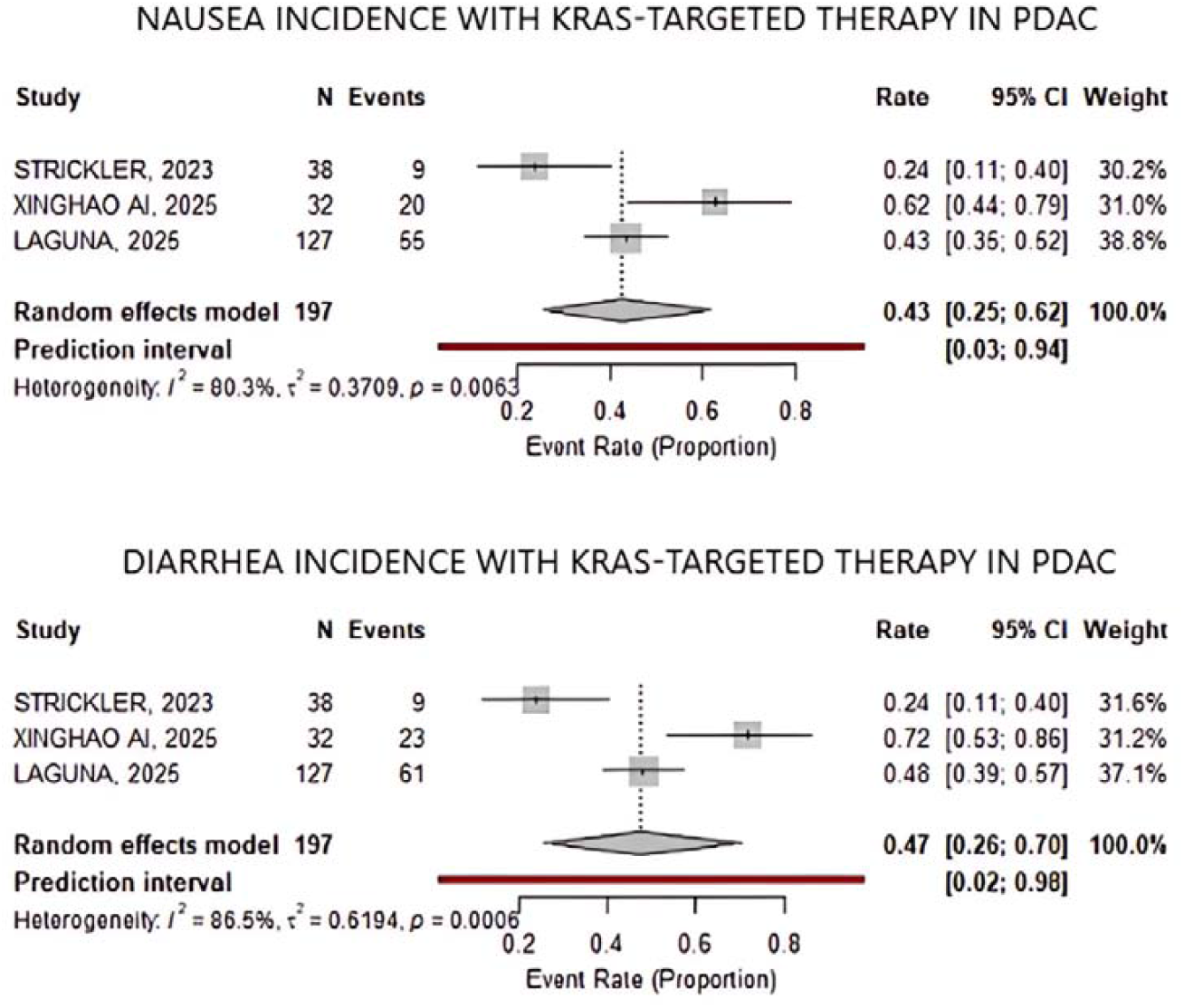
Incidence of gastrointestinal adverse events with KRAS-targeted therapy in PDAC. Pooled analysis of three trials (n = 197) showed nausea and diarrhea incidences of 43% (95% CI, 25–62%) and 47% (95% CI, 26–70%), respectively, with wide prediction intervals and moderate to substantial heterogeneity (I^2^ = 80.3% and 86.5%).

The risk of bias was substantial. The ROBINS-I assessments indicated a moderate-to-high risk of bias across domains, including confounding, selection, and outcome reporting. In the absence of control arms, response rates cannot be reliably contextualized against the natural history of PDAC [Supplementary Appendix]. Funnel plot analysis revealed no evidence of publication bias. The studies were largely symmetrically distributed around the pooled estimate, with greater dispersion among smaller trials, as expected [**Figure 4**].

**Figure 4:**
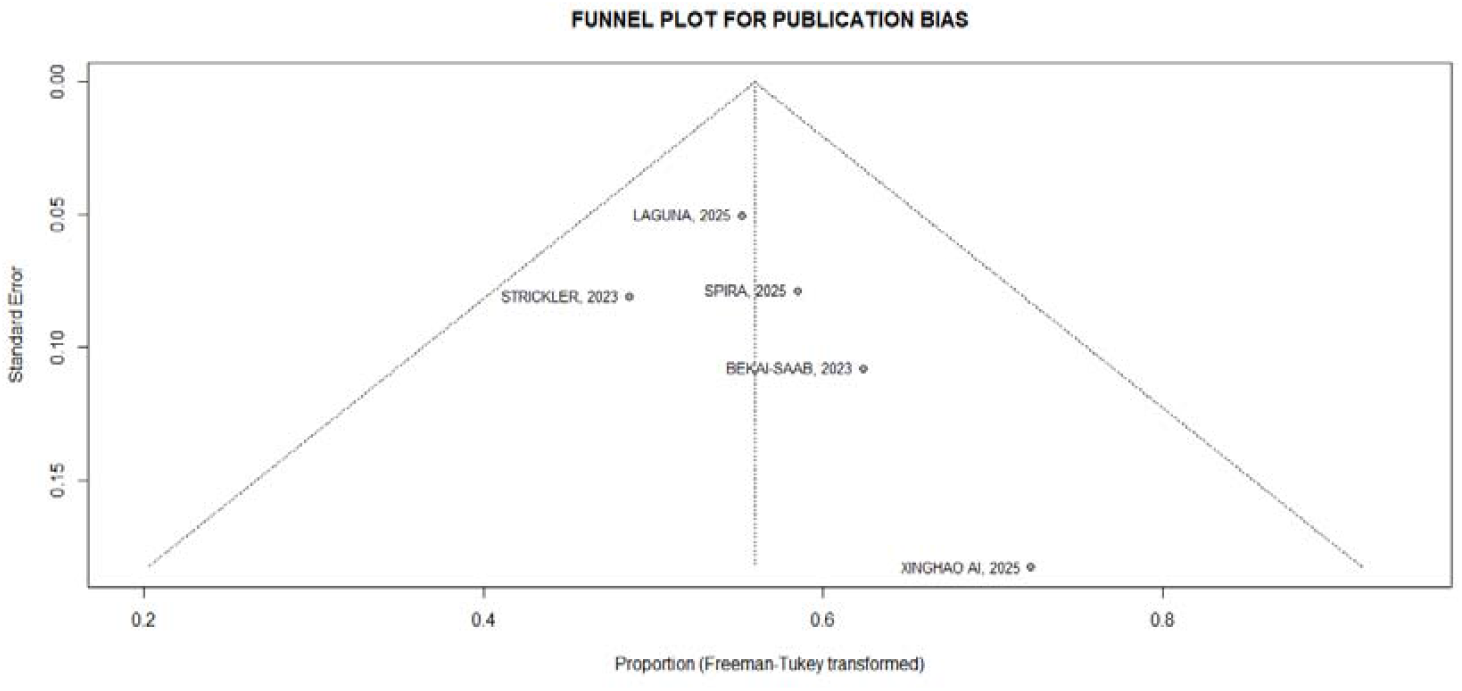
Funnel plot of the included studies. The plot appears largely symmetrical, although one smaller study in the lower right quadrant showed a higher response rate than the pooled estimate, suggesting mild asymmetry but no clear evidence of publication bias.

Taken together, the results of this meta-analysis indicate that direct KRAS inhibition in PDAC is both feasible and biologically active, although the supporting clinical evidence is limited, heterogeneous, and prone to bias, warranting cautious interpretation of the results. Nonetheless, these studies contribute to defining an evolving therapeutic landscape. After decades in which KRAS was deemed “undruggable,” the emergence of covalent inhibitors and mutation-directed delivery systems represents a paradigm shift. The modest but measurable activity validates the biological rationale and establishes a foundation for further development. Moving forward, progress will hinge on allele-specific inhibitors, rational combinations with immunotherapy and targeted agents, mutation-directed vaccines, and, critically, the standardization of trial design, endpoints, and biomarker integration, such as ctDNA dynamics and resistance profiling, to achieve durable benefits.

## Methods

### Search strategy and study selection

We performed a systematic review and meta-analysis to evaluate the efficacy and safety of KRAS inhibitors in pancreatic ductal adenocarcinoma (PDAC). A comprehensive literature search was conducted in MEDLINE (via PubMed), Embase, and the Cochrane Library using predefined Medical Subject Headings (MeSH) and free-text keywords. The search terms included: (“pancreatic neoplasms” OR “carcinoma, pancreatic ductal” OR “pancreatic” [tiab]) AND (“Proto-Oncogene Proteins p21(ras)” OR “ras proteins” OR KRAS OR G12D OR GFH375 OR G12C) AND (Sotorasib OR Adagrasib OR Pyridines OR Krazati OR Piperazines OR MRTX849 OR “RNAi therapeutics” OR Daraxonrasib). The search was last updated in 2025. The reference lists of relevant articles were manually screened to identify additional eligible studies. Full-text publications and meeting abstracts were included.

### Eligibility criteria

Studies were included if they met the following criteria: (1) enrolled patients with KRAS-mutated PDAC; (2) included patients with previously treated metastatic or locally advanced disease; and (3) evaluated the use of KRAS inhibitors as second-line therapy. The exclusion criteria were as follows: (1) overlapping patient populations across reports, (2) use of KRAS inhibitors as first-line therapy, and (3) studies on non-pancreatic primary tumors.

### Study selection and data extraction

Two independent reviewers (K.O.M.T. and O.C.) screened the titles, abstracts, and full texts against predefined criteria. Disagreements were resolved through discussion and consensus. Data were extracted from each eligible study on the objective response rate (ORR), defined as partial or complete response, and safety outcomes, focusing on the most frequently reported adverse events.

### Risk of bias and publication bias

The risk of bias in non-randomized studies was assessed using the ROBINS-I tool. Publication bias was evaluated by visually inspecting the funnel plot, and the Freeman-Tukey double arcsine transformation was used to stabilize the variance of the proportions before calculating the pooled effect.

### Statistical analysis

All statistical analyses were performed using R software (version 4.5.1; R Foundation for Statistical Computing, 2025) in accordance with the PRISMA guidelines. Patients were dichotomized as those who achieved or did not achieve ORR. A single-arm meta-analysis of proportions was conducted to estimate the pooled ORR and incidence of adverse events, each with corresponding 95% confidence intervals (CI). Between-study heterogeneity was assessed using Cochran’s Q test and quantified with the I^2^ statistic, with thresholds of I^2^ >25% or p <0.10 indicating significant heterogeneity. A random-effects model was applied unless significant heterogeneity warranted the use of a random-effects model. Sensitivity analyses were performed to assess the robustness of these findings.

## Data Availability

The data generated and analyzed during this study are available from the corresponding author upon reasonable requests.

## Author contributions

All authors contributed to study conception, design, data collection, analysis, and manuscript preparation.

