## Supplementary for "KRAS Inhibition in Pancreatic Cancer: Pooled Efficacy and Safety Signals from Early-Phase Studies": Supplementary.pdf

**Supplementary Figure X. PRISMA flow diagram of study selection process:**

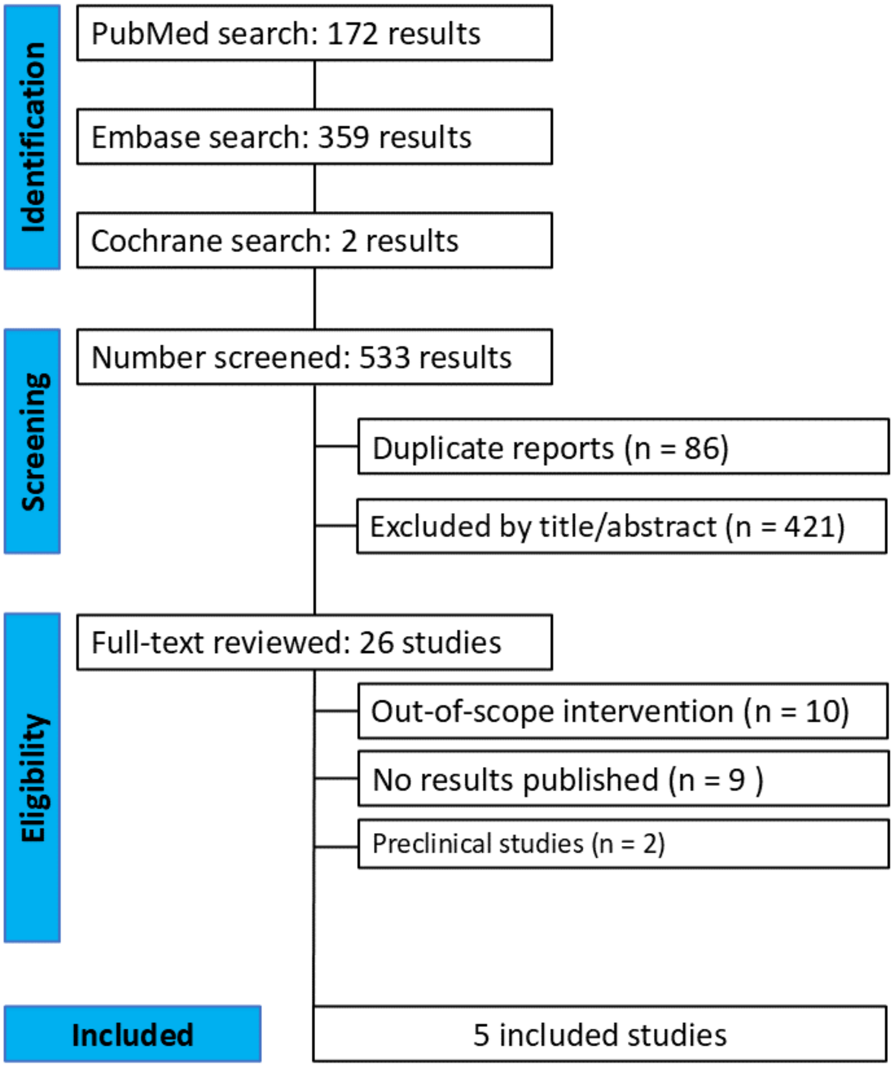


Figure 1. PRISMA flow diagram of study screening and selection

***Figure 1.*** *PRISMA flow diagram of study screening and selection. The systematic search identified 533 records from PubMed, Embase, and Cochrane databases. After removal of duplicates and exclusion by title/abstract screening, 26 studies were assessed for eligibility. Of these, 10 were excluded due to out-of-scope interventions, 9 had no published results, and 2 were preclinical studies. Five studies met the inclusion criteria and were included in the meta-analysis.*

**Table S1. Risk of bias assessment across domains (ROBINS-I). Visualization created using the robvis tool (McGuinness & Higgins, 2020).**

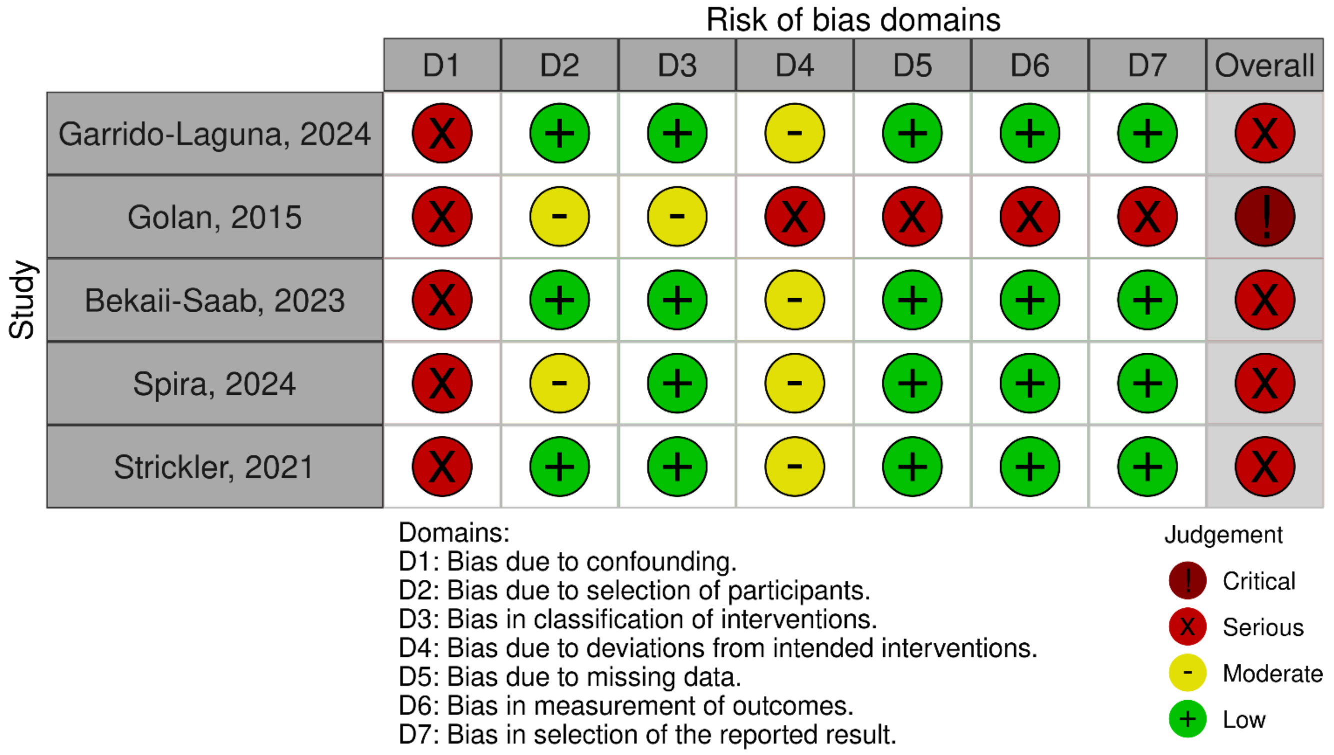


*D1: Bias due to confounding; D2: Bias in selection of participants into the study; D3: Bias in classification of interventions; D4: Bias due to deviations from intended interventions; D5: Bias due to missing data; D6: Bias in measurement of outcomes; D7: Bias in selection of the reported result. Symbols: green (+) = low risk; yellow (–) = moderate risk; red (×) = high risk; exclamation (!) = critical risk.*

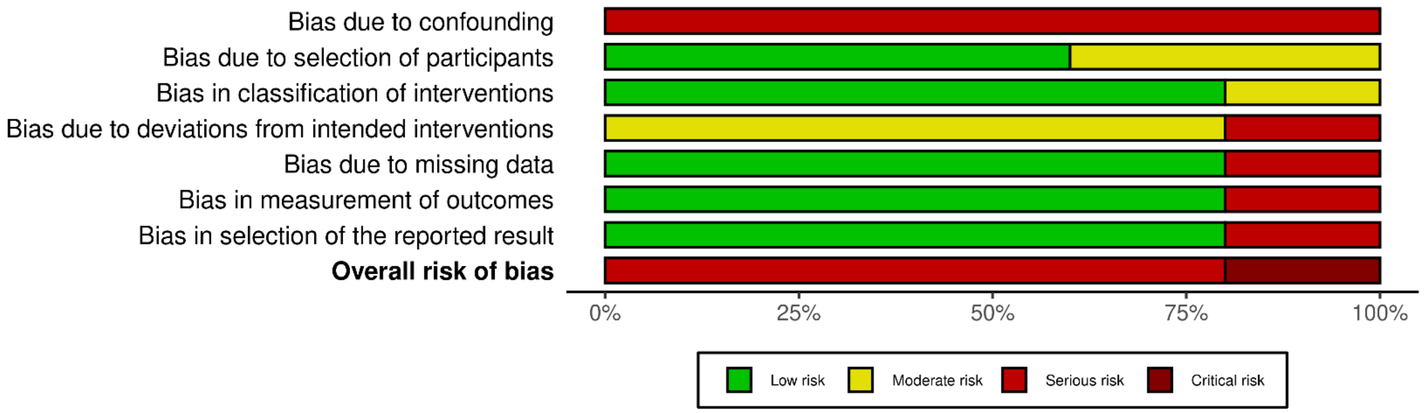
